# Graph connection Laplacian allows for enhanced outcomes of consumer camera based photoplethysmography imaging

**DOI:** 10.1101/2024.03.07.24303826

**Authors:** Stefan Borik, Hau-Tieng Wu, Kirk H. Shelley, Aymen A. Alian

**Affiliations:** Dept. of Anesthesiology, Yale University, New Haven, CT, 06510, USA; Dept. of Electromagnetic and Biomedical Eng., University of Zilina, Zilina, 01026, Slovakia; Courant Institute of Mathematical Sciences, New York University, New York, 10012, USA

## Abstract

**Object:** This work introduces a novel method to minimize the effect of global phase deviation that is inherent in photoplethysmographic images (PPGI) captured by video.

**Method:** We analyzed the facial vascular network obtained from a consumer camera as a two-dimensional manifold. Subtle phase variations across skin sites are due to complex dynamics of the vascular tree. Utilizing PPGI, the phase is modeled as a vector field of the facial manifold. The phase variations over different skin sites are caused by different blood volume modulations. We propose using the Graph Connection Laplacian (GCL) technique to quantify the global phase deviation, with the hope that correcting for this deviation could improve the quality of the PPGI signal. It is also postulated that study of this phase deviation might reveal valuable anatomical and physiological information.

**Result:** The proposed algorithm appears to yield a higher-quality global PPGI signal. By correcting the global phase deviation estimated by GCL waveform features such as the dicrotic notch are emphasized. The perfusion map, with the global phase deviation (estimated by GCL as intensity), appears to reflect skin perfusion dynamics.

**Conclusion:** This algorithm enhances the quality of the global PPGI signal, facilitating the analysis of morphological parameters and showing promise for advancing PPGI applications in scientific research and clinical practice.

## 1. Introduction

The landscape of cardiovascular health monitoring has undergone a notable transformation with the introduction of smartphone cameras. This technology allows for extraction and analysis of photoplethysmography (PPG) signals. Camera-based PPG imaging (PPGI) has evolved from the original contact-based methods which trace their origin back to the early 20th century [1]. Initially developed to measure blood volume changes in the microvascular bed, PPG has evolved over time to incorporate imaging modalities, providing a non-invasive means of assessing physiological parameters [2]. The advent of consumer electronics and advanced imaging technologies (like CCD earlier [3] and now CMOS sensors [4]) has propelled PPGI into the spotlight, particularly in the last two decades, starting with pioneering works of Blazek et al. [3], [5]. The PPGI typically involves capturing variations in light absorption or reflection at the skin surface, primarily driven by pulsatile blood flow changes. This technique has found diverse applications, ranging from pulse oximetry [6], [7], [8], [9] and cardiovascular monitoring to more recent innovations in facial PPG for health assessments [10], [11]. The ability of PPGI to provide contactless measurements has gathered interest also in another fields such as vital sign monitoring [12], [13], [14], [15], [16], wound assessment [17], [18], driver state estimation [19], [20], [21] or pain evaluation [22]. The continuous refinement of PPGI methodologies and the exploration of novel applications underscore its significance in modern healthcare and wellness monitoring.

Researchers and practitioners are increasingly turning to additional techniques to broaden their analysis and to push the boundaries of PPGI capabilities. Starting with traditional spatial averaging [12], which primarily aims to increase the signal-to-noise ratio, different methods are now being used. They differ in complexity or approach to signal extraction. Some of these include the R-G algorithm [23] (combining the red and green channels in the case of PPGI via an RGB camera), and the POS (Plane-Orthogonal-to-Skin) algorithm [24], (which works with all three layers). Others such as the historically sorted G [12], (which works only with the green layer or greyscale data), G-R [23], PCA [25], ICA [26], CHROM [27], PBV [28], 2SR [29], synchrosqueezing transform [30], the aforementioned POS [24] or Face2PPG [31]. A new approach is deep or machine learning methods [32], [33], which bring additional potential for feature extraction, classification and understanding of the complex links within the PPG signal. Also worth mentioning, is using a lock in amplification method with the reference PPG signal either from an external device or by the averaging of skin pixels from whole recorded area [34], [35].

There is an unavoidable global phase (or temporal) deviation across a body of tissue caused by the properties of the vascular tree. This is a system of interconnecting and branching elastic vessels where the pulse waves (i.e. direct and backward) propagate in different directions and velocities from the heart towards the periphery and back. Further complication is caused by the interaction of this vascular tree with the autonomic system causing both localized and global changes in vascular tone. The main novelty of this paper is to quantify and modify the global phase deviation based upon the Graph Connection Laplacian (GCL) [36], [37] method of synchronization. Phase deviations in PPGI signals at different skin sites are intricately linked to the dynamic blood flow pattern in the vascular network. Our hypothesis is that this hemodynamic-induced phase variation embodies valuable anatomical information and potentially other relevant details of interest. We model the vascular network underlying the facial pixels (which in this case are recorded using a consumer camera) as a two-dimensional manifold that encodes the phase of localized blood flow. The goal is to use PPGI over each pixel to reconstruct this phase information and thus obtain an integral and improved global PPGI curve. This could then be used to analyze the morphological parameters and also improve the spatial resolution of PPGI in mapping skin perfusion dynamics.

## 2. Materials and Methods

In this section, we provide a detailed description of our algorithm (the overall performance of the algorithm is shown in Fig. 1, and the flow chart of the whole algorithm is shown in Fig. 2), which aims to extract tissue perfusion information from video recording acquired by consumer electronics (a smartphone camera) and improve the properties of the detected PPG curve in terms of its morphology and the detection of spatial dynamics across the recorded region.

**Fig. 1:**
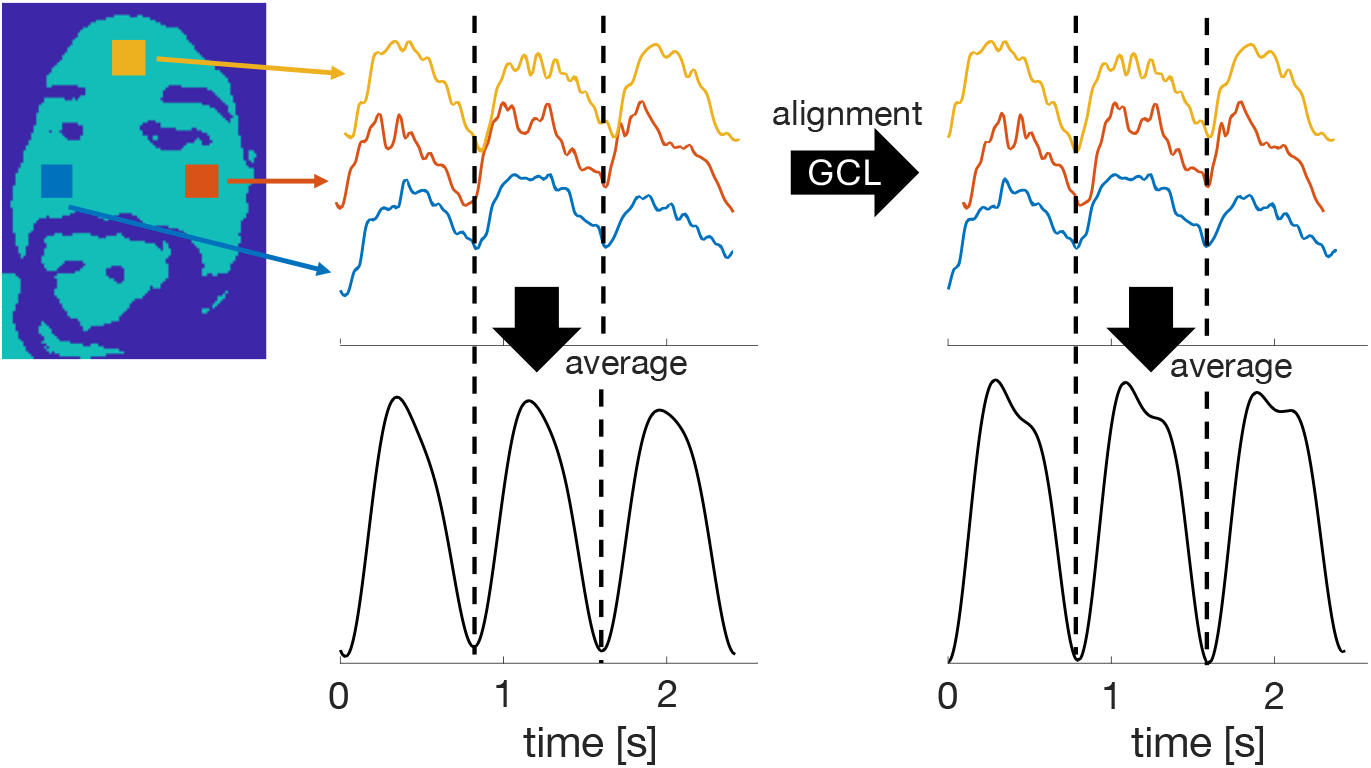
Illustration of the signal alignment from different measurement sites and its impact on the global PPGI waveform when averaged all face channels once averaged without GCL (left) and with GCL (right).

**Fig. 2:**
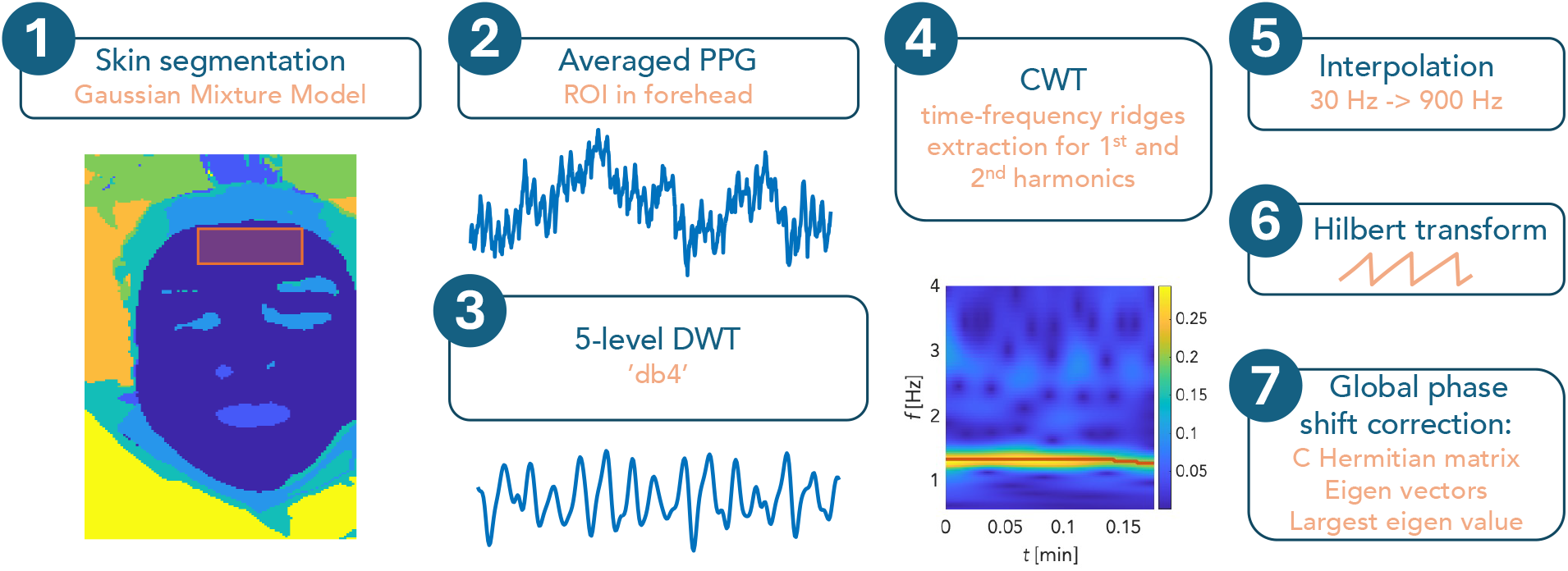
The overall algorithm flowchart.

### 2.1. Experimental setup

The experimental setup is based on a study conducted on volunteers undergoing a lower body negative pressure (LBNP) procedure [38], with their faces being recorded using a consumer-quality camera, namely the iPhone 11 (Apple, Cupertino, CA, USA). During recording, automatic settings such as autofocus were turned off while exposure was locked to prevent changes in the image that could reduce the quality of the perfusion information. Video was recorded in H264 compression format with a frame rate of 30 Hz. The subject’s face in supine position was illuminated with white diffuse LED light. As part of the comprehensive LBNP protocol, we selected only a 30 s segment from the baseline portion of the recording. The study protocol was approved by the Institutional Review Board of Yale University (No. 2000031899). Informed consent was obtained from all subjects prior to the study. All methods were performed in accordance with the study protocol and with the Declaration of Helsinki. To verify the functionality and assess the performance of the algorithm, we selected 20 subjects (10 females and 10 males) aged 32.18±3.36 yrs.

### 2.2. Photoplethysmography signal extraction from video

The first step was the extraction of PPG signals from the video recording. In this respect, the camera sensor can be thought of as a PPG sensor matrix, where in theory each pixel of the image sequence can carry spatial information about the blood supply of a given location. The SNR of consumer cameras is usually not sufficient to extract the PPG signal from only one pixel, and different techniques have been used to increase the SNR. In most cases, the first step is spatial averaging of pixels either in the form of spatial down sampling [9] or through a moving kernel with the desired overlap [39]. In our case, we average the pixel values in a box of size 30×30 px^2^ with 10 px overlap which represents cumulating pixel information from the areas of approximately 1 cm^2^ at given camera distance and its field of view. We call an overlapping region of size 30×30 pixels a channel and suppose we obtain *L* channels. This step will not only increase the SNR of the extracted PPGI signals, but also reduce the number of signals to work with while still maintain a reasonable spatial resolution. The result of this process (see Fig. 3) is a raw PPG signal that we decompose into individual color layers, including red (R), green (G), and blue (B), and store on disk for further analysis. Next, we work only with the green (G) channel, which contains the most information about perfusion. This fact is related to the technical realization of the color camera (Bayer mask), the wavelength of the green light and its penetration into the tissue, as well as the absorption and scattering properties of the skin. The main luminophore in this case is blood hemoglobin [39], [40], [41]. Consequently, we denote each raw PPGI signal collected from the *i*^th^ channel as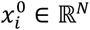, where *N* = 900 is the number of sampling points over the 30 s segment with the frame rate of 30 Hz. The pseudo code for the process related to this paragraph is shown in Algorithm 1.

**Fig. 3:**
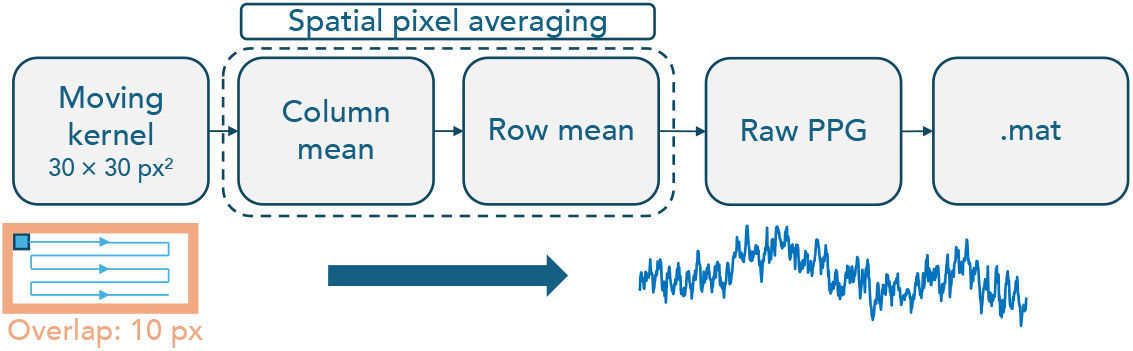
Spatial averaging and obtaining PPGI signals

#### Algorithm 1.

Spatial averaging and obtaining PPGI channels.

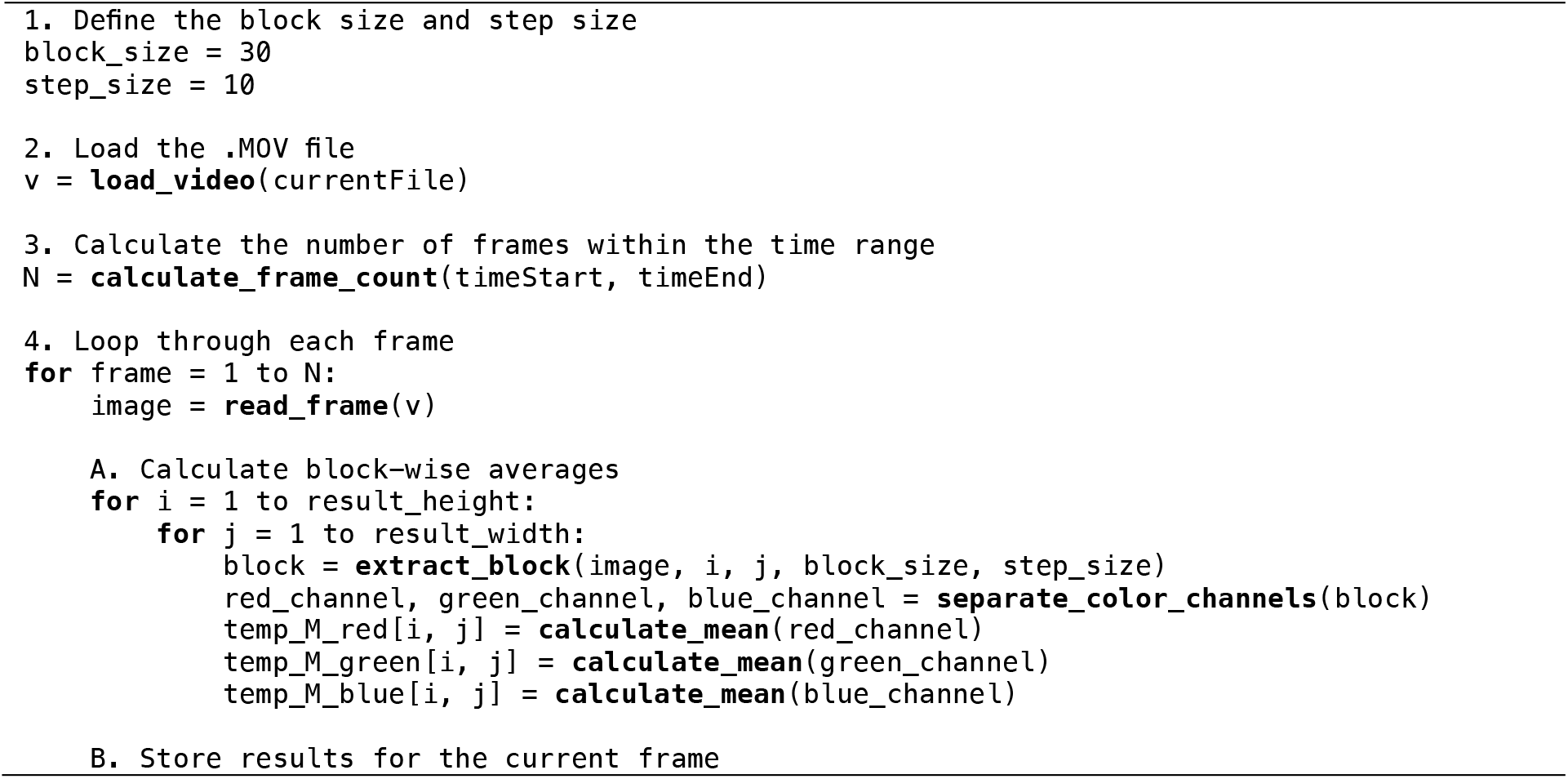
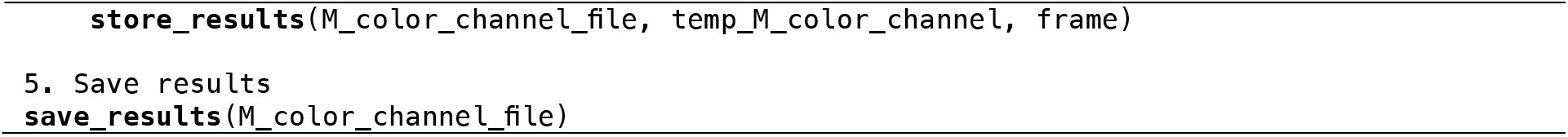

### 2.3. Signal pre-processing and filtering

Further signal processing proceeds by segmenting the skin using a Gaussian Mixture Model (GMM) to ensure that we continue to work only with living skin pixels where we can expect PPG signals to be present (see the dark blue region in Figure 2(a)). The segmentation is semi-automatic and allows the user to control the detected clusters and add them as needed.

The next step involves a discrete wavelet transform (DWT) for each PPGI channel to select a band with the range 0.4 – 4 Hz that we can assume cardiac activity. For extraction of the signals related to cardiac activity, we use the Daubechies 4 wavelet (db4), with the number of DWT levels/octaves set to 5, and denote the resulting signals as 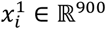.

After the DWT, we select a segment with minimal motion artifact in the following way. First, we find all channels associated with the subject’s forehead region (see the red box/ROI in Figure 2(a)) and obtain average of the associated signals, which is denoted as 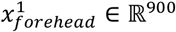. From 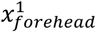. Subsequently, we manually extract a high quality ten-second interval that is less impacted by motion artifacts. Then, we restrict 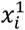 on the selected ten-second interval and denote the resulting signals as 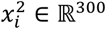.

The process continues by extracting the first and second harmonics of each PPGI signal 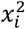 based on the continuous wavelet transform (CWT) using the time-frequency ridge method, followed by the inverse CWT and interpolating from the original 30 Hz to 900 Hz in order to retrieve subtle phase shifts across the signals, which is denoted as *x*_*i*_ ∈ ℝ^9,000^. Since the PPGI signals are generally noisy and we use relatively small averaging regions to keep the spatial information, the second harmonic component in the CWT spectrum is usually overlaid with noise. To extract it, we use the frequency ridge by simply considering doubling the extracted ridge of the first harmonic component, and then extract the second harmonic component. The first and second harmonics of the *i*th channel are subsequently denoted as *h*_1,*i*_ ∈ ℝ^9,000^ and *h*_2,*i*_ ∈ ℝ^9,000^ respectively.

Next, the signal is trimmed to the length of only 3 consecutive heartbeats, taking into account both the phase alignment dependence of the data length (influenced by heart rate variability) and the exact selection starting from the peak or valley of the PPG curve. A Hilbert transform is then performed. The pseudo code for the process related to this paragraph is shown in Algorithm 2.

#### Algorithm 2.

Signal pre-processing and filtering.

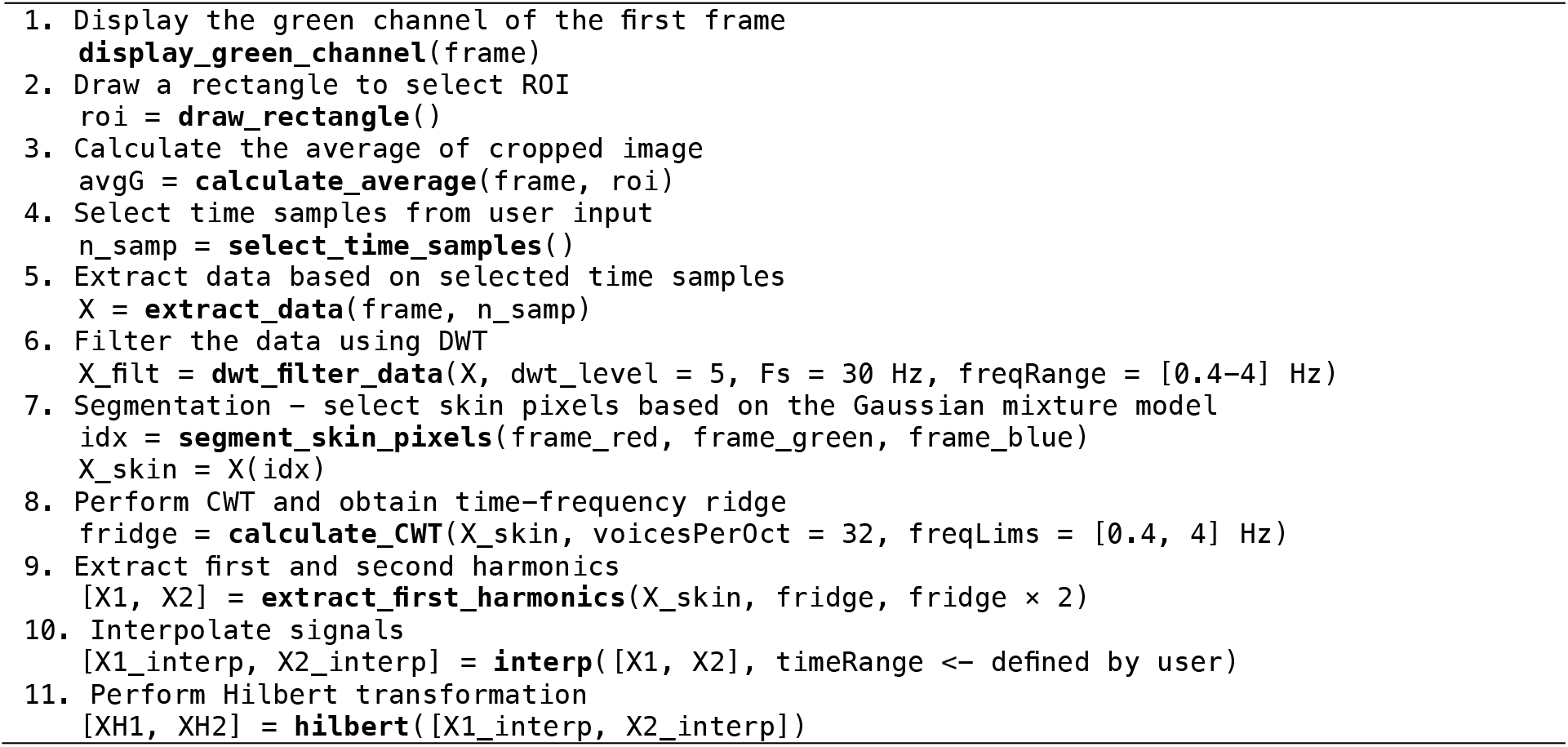

### 2.4. Synchronization of the extracted PPG signals

Due to the inevitable global phase deviation caused by hemodynamics, the main novelty of this paper is applying the GCL [36], [37] to adjust the global phase deviation. In a physiological context, the phase variations in PPGI across different pixels are intricately linked to the dynamic progression of blood flow within the vascular network. Our hypothesis posits that this phase deviation induced by hemodynamics encapsulates valuable anatomical and physiological information and potentially other pertinent details of interest. We model the vascular network underlying the facial pixels as a 2-dim manifold and the geometric structure encodes the phase of blood flow changes. The goal is to use the PPGI sensors over each pixel to recreate this phase information and hence the vascular network structure. We first estimate the pairwise phase deviation between any two PPGI channels *x*_*i*_ and *x*_*j*_ using their 1^st^ harmonics *h*_1,*i*_ and *h*_1,*j*_ and compute:

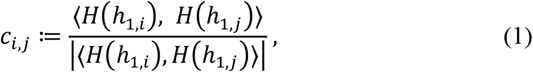

where *H* converts the real-form signal into its analytic companion, defined as 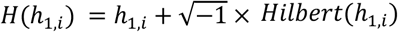, with *Hilbert* the Hilbert transform, and ⟨, ⟩ is the inner product of two complex signals. The entry *c*_*i,j*_ estimates the global phase shift of *x*_*i*_ and *x*_*j*_. See [38] for more discussion about the nonlinear relationship between harmonic phase and PPG morphology and Discussion for more technical details. Since we have *L* PPGI channels, we obtain a Hermitian function *C* of size *L* × *L*, called the graph connection Laplacian (GCL), so that the (*i, j*)^th^ entry of *C* is *c*_*i,j*_ if the ith channel and jth channel are within 5 cm distance, and 0 otherwise. We find the eigenvector of *C*, denoted as *w*_1,_ *w*_2_ …, with the associated eigenvalues ranked from large too small. Then, the global phase deviation of the *i*^th^ PPGI channel is then corrected by:

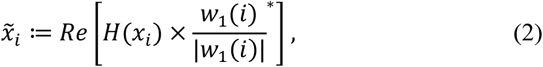

where *Re* means taking the real part and the superscript * means taking the complex conjugation. The philosophy underlying this correction is that *w*_1_(*i*) recovers the blood flow phase underlying the *i*^th^ channel. In Fig. 4, an illustration of synchronization with GCL when there are only two channels is shown. See discussion section for more technical details of this GCL algorithm. The pseudo code for the process related to this paragraph is shown in Algorithm 3.

**Fig. 4:**
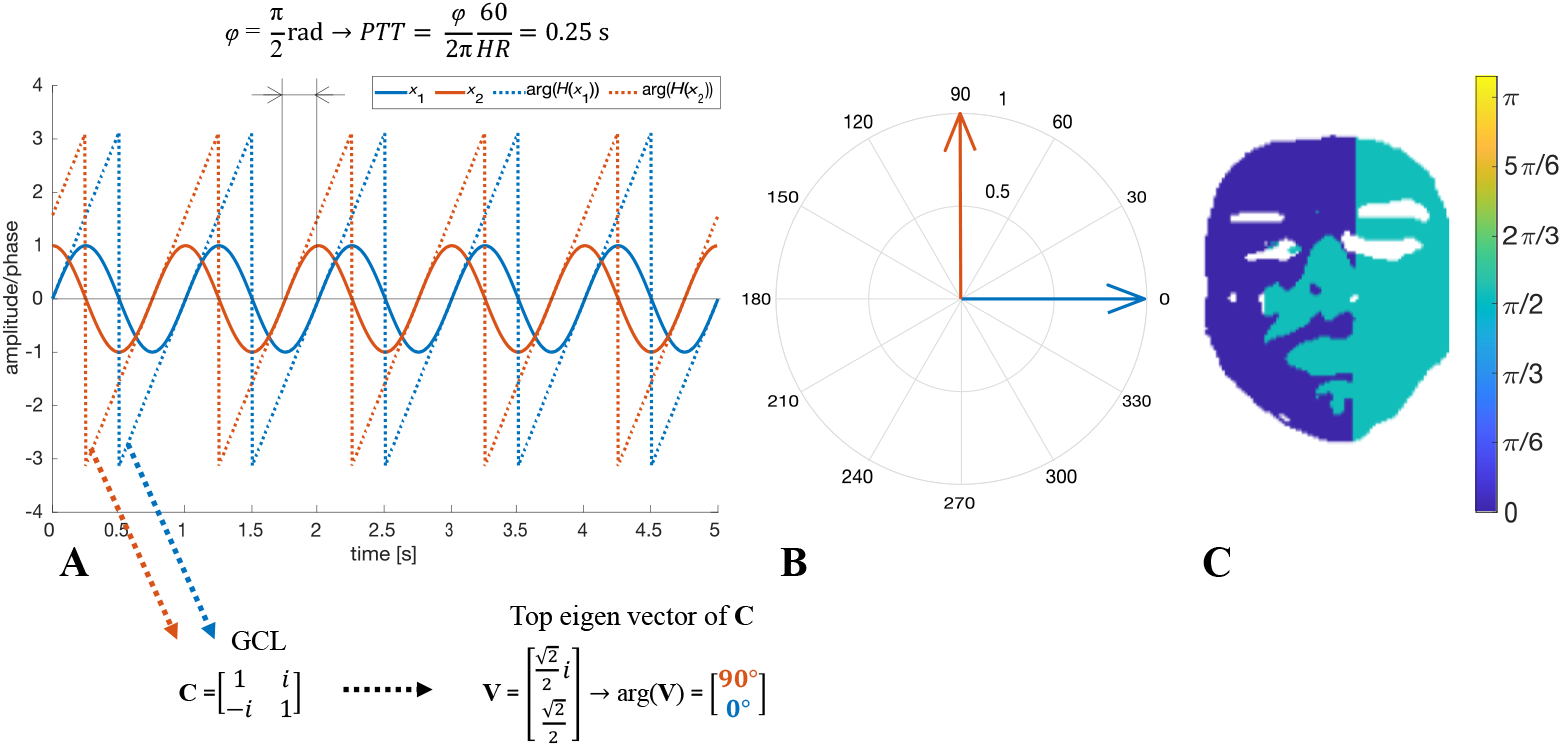
Illustration of the GCL-based synchronization. A: Two sine waves, *x*_1_=sin(*ωt*) and *x*_2_=sin(*ωt+* π/2), where *ω* = 2π with a global phase shift of π/2, where we assume *x*_1_ is recorded from the left hand side of a simulated face shown in C and *x*_2_ is from the right hand side. Dotted lines depict phases of the signals determined by applying Hilbert transformation to *x*_1_ and ***x***_**2**_. The phases are then used to construct GCL matrix. After finding the top eigenvector of the GCL matrix, we obtain the phase of the first sine wave, shown in the red arrow in (B), and the phase of the second sine wave in the blue arrow. In addition, we show how the phase shift is related to pulse transit time (*PTT)*. In this simulated case, the phase shift of π/2 rad between *x*_1_ and *x*_2_ is related to *PTT* = 0.25s.

#### Algorithm 3.

Phase shift estimation and synchronization.

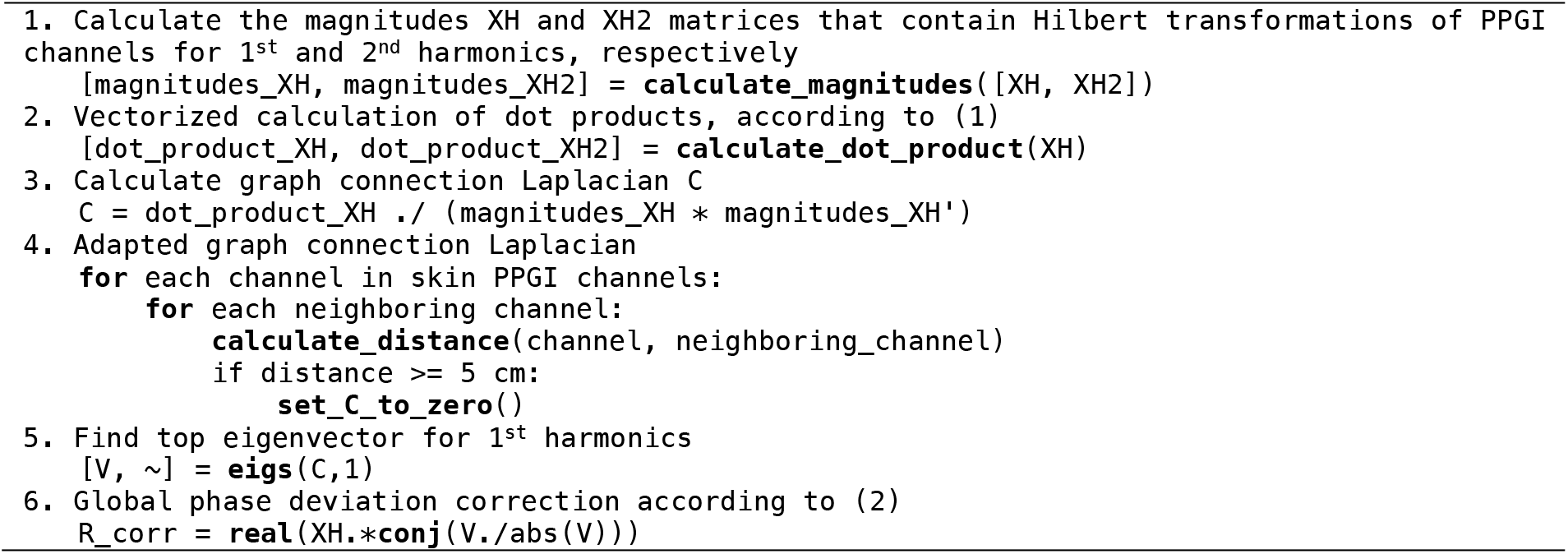

### 2.5. Construction of the final non-contact PPG signal

We construct several non-contact PPG signals. The first one is by a simple averaging of all unaligned PPGI channels; that is, 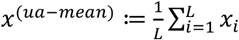. The second one is averaging all aligned PPGI channels; that is 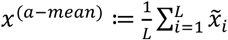.

### 2.6. Construction of perfusion maps

We propose the following approach to visualize perfusion patterns in the face. We generate images encoding the energy of the first harmonic, denoted as *I*_*E*,1_, which is defined as the energy of the 1^st^ harmonic of each channel. We call these maps *energy maps*.

Another set mapping the skin perfusion properties is generated from the top eigenvector of the GCL, *w*_1_. The phase encoded in 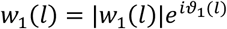, reflects the phase shift of the *l* th channel over the face. Thus, by generating a color image of the face region with the color in each pixel reflecting the entry *ϑ*_1_, we obtain the resulting images, which we denote as *I*_*phase*_, and we call these maps as *phase maps*.

For each image, to avoid the impact of oversaturation, we replace all values larger than 95% percentile by the 95% percentile before plotting. For phase maps, we also trim colormap based on the range of median phase ±1rad of entire skin area in order to highlight subtle difference in phase shifts across the PPGI channels.

## 3. Statistical analysis

To quantify the performance of phase synchronization for PPGI signal 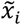, we consider the following metric. For the raw unaligned signals, we apply the Hilbert transform and obtain the complex form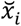. We then convert the amplitude and phase into the phasor form at the center point:

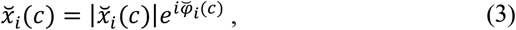

where *c* denotes the center point that we extract the phasor and 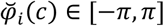. For the aligned signals, which are the output of (2), we apply the Hilbert transform to obtain the complex form 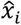. We then convert the amplitude and phase into the phasor form at the center point:

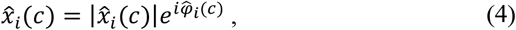

where *c* denotes the center point that we extract the phasor and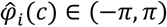. In addition, we denote the median of 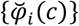 as 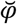 and denote the median of 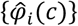 as 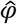. To check if 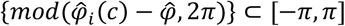 and 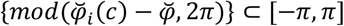 have the same distribution, we apply the Kolmogorov-Smirnov test with p<0.05 viewed as statistically significant. Also, we apply F-test to check if 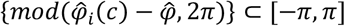 and 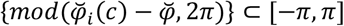 have the same variance with p<0.05 viewed as statistically significant. We also performed Wilcoxon signed-rank test to investigate if the phase shift on the cheeks and forehead, where the phase deviation of the *i*th channel is quantified by the angle of 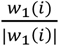, are different with p<0.05 viewed as statistically significant.

Furthermore, we conduct a statistical analysis to examine the impact of synchronization on the generation of the final non-contact PPG signal. We extract the amplitudes of the harmonic components from the spectrum of the final non-contact PPG signals, one with and one without GCL synchronization. Applying the left-tailed Wilcoxon signed-rank test, we test the null hypothesis that GCL synchronization results in higher amplitudes of the first and second harmonics. A p-value less than 0.05 is considered statistically significant, and we employ Bonferroni correction to address multiple testing.

## 4. Results

The first step to verify the quality of the phase alignment of the PPGI curves was to plot the phasors separately for 1^st^ harmonic component. The distribution of phasors of all PPGIs channels evaluated by (6) of all subjects is shown in Fig. 5, where we plot 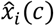 as a vector, with the magnitude 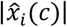 and angle *φ*_*i*_(*c*). It can also be noticed in Fig. 5 that the unaligned phasors (blue arrows) tend to be directed and arranged in two or more clusters for some subjects, while aligned phasors are in general well clustered in one direction. The Kolmogorov-Smirnov test showed that the two distributions are significantly different with *p <* 10^−4^, and the F-test showed that the two distributions have significantly different variance with *p <* 10^−4^. On the other hand, we shall emphasize the observation that dominant direction 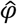 of aligned PPGI signals differs from subject to subject. This is the consequence of the nonuniqueness of GCL approach inherited from the freedom of eigendecomposition.

**Fig. 5:**
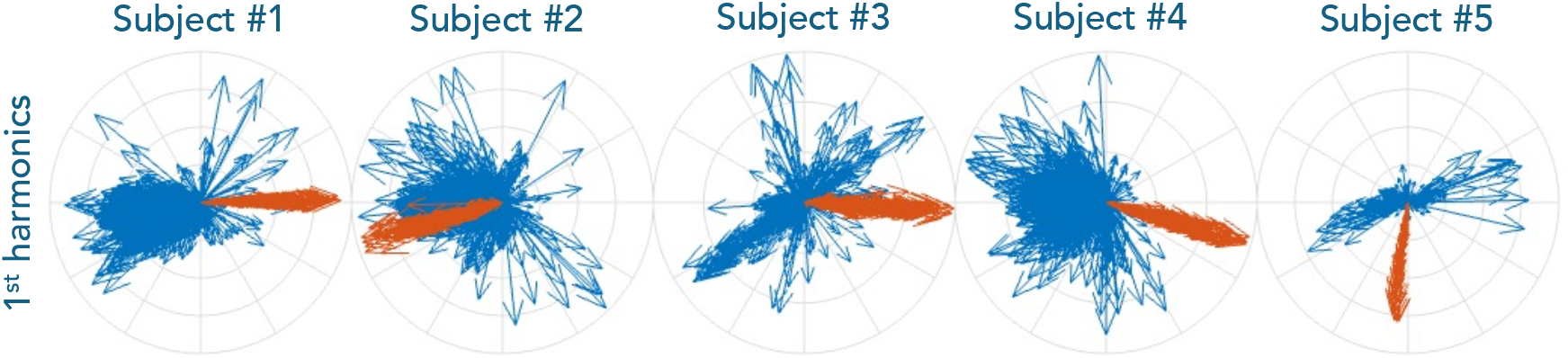
Visualizaton of phase alignment. Five compass plots in the top row depict arrows representing aligned (red) and unaligned (blue) states across subjects, where columns represent different subjects. In the bottom row, the alignment is carried out using the second harmonic.

After aligning the PPGI signals and verifying the quality of their alignment, we obtain various final non-contact PPG signals, including *x*^(*ua*-*mean*)^, *x*^(*a*-*mean*)^ shown in Fig. 6. One of the interesting effects of phase alignment is the appearance of the dicrotic notch, which is most pronounced for Subject 2 in Fig. 6. Another observation is that the average of phase aligned PPGIs contributes to maximizing the amplitude of the PPGI signal.

**Fig. 6:**
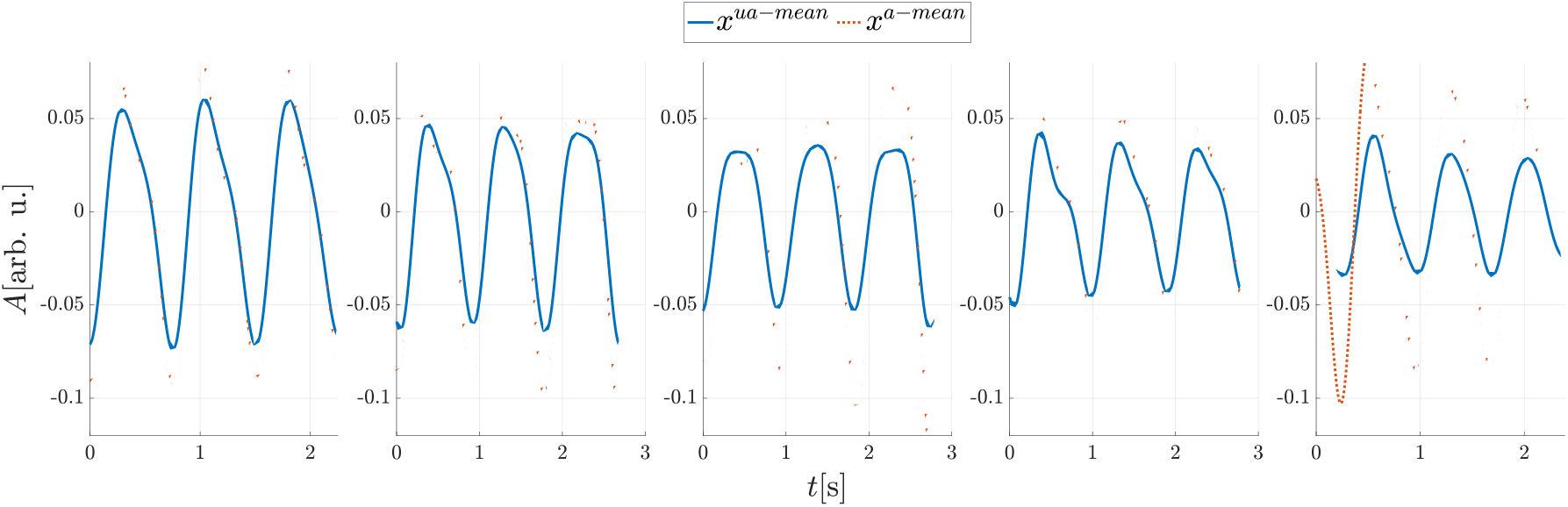
Comparison of averaging phase unaligned and phase aligned PPGI waveforms from all channels to generate the final non-contact PPG signal. Solid blue curves are from the unaligned PPGI signals, while dashed red curves are from the aligned PPGI signals. From left to right: results from subjects 1 to 5 respectively.

To quantify these findings, we conduct the left-tailed Wilcoxon signed-rank test on all 20 subjects. This test aims to determine whether the amplitudes of the first and second harmonics of the final non-contact PPG signal are higher when GCL synchronization is applied. The results of this statistical analysis demonstrate that GCL synchronization leads to significantly higher amplitudes of both the first and second harmonic components, with p-values less than 0.001 for both harmonics.

Next, we demonstrate the perfusion maps derived from PPGI. See Fig. 7 and Fig. 8 for our 5 different subjects, who were selected based on specific characteristics such as skin tone, facial hair, facial mask or extensive makeup. We will pay specific attention to the difference between cheeks and forehead, the two dominant areas on the face that are supplied by different arteries.

**Fig. 7:** Results of the two perfusion maps for the first three subjects with diverse characteristics, including skin tone, facial hair, mask. For each subject, the left subfigure is the energy map, and the right subfigure is the phase map. **Figures are not shown due to the Medrxiv’s policy**.

**Fig. 8:** Results of the two perfusion maps for the last two subjects with diverse characteristics, including skin tone and makeup. For each subject, the left subfigure is the energy map, and the right subfigure is the phase map. **Figures are not shown due to the Medrxiv’s policy**.

For subject 1 we can observe different captured amplitudes in the cheeks and glabella region in the energy maps, which might be interpreted as ‘‘cheeks are more perfused’’. Another interesting observation for subject 1 is its phase map associated with *w*_1_, where we can see clustering at specific locations in the image. There is a slightly different phase shift in the right side of the face. The forehead area also exhibits different phase shift compared to other parts of the face which could be related to different arterial supply.

Subject 2 is specific to his facial hair (chin and beard), which may be a source of motion artifacts that may deceptively influence the detected PPGI signal amplitude. Again, the cheeks have more dominant energy compared to forehead also in this case. In the phase map, while it is less obvious, the phase in the forehead is different from that in the cheek.

Subject 3 is specific in that he is wearing a face shroud and also has goggles on his eyes. For this case, it is not possible to obtain information over cheek, so a comparison between cheeks and forehead is not possible.

For subject 4 (Fig. 8), we notice a distinct vertical line across the middle of the forehead, which may be a wrinkle or an ongoing supratrochlear vein. This area is even more pronounced in the phase map. Again, we can see a phase difference between the forehead and cheeks in the phase map.

We can also see an interesting result for subject 5 (Fig. 8), where the glowing eye surroundings can be tracked in the energy maps. In this case, this is the result of different makeup on the face and around the eyes. In this case, this is a consequence of the different makeup on the face and around the eyes. In this case, the phase difference between the forehead and cheeks is less clear in the phase map.

Wilcoxon signed-rank test showed that the phase deviation is significantly different in forehead and cheeks for subject 1, 2 and 4 (with *p* < 10^−4^). We did not test subject 2 because we cannot access the cheeks due to the facial mask. We also omitted subject 5 due the extensive makeup.

## 5. Discussion and Conclusion

Our aim is to advance the signal processing techniques for the PPGI in two ways. First we apply the GCL-based synchronization algorithm to construct a better-quality global PPG. This higher quality PPG waveform contains details that are more challenging to obtain using traditional approaches. Examples of these details include the dicrotic notch and its precisely localized position. The novelty here is to view GCL as a “coupler” for all the PPGI signals in the studied area. Then we can determine the phase discrepancy that arises from the pulse wave as it travels through the vascular tree. Using this information, we can synchronize the PPGI signal across the image. The results show that this unique method can enhance the features of the PPG waveform (see Fig. 6) and boosts its amplitude. The second approach introduces the use of novel images called perfusion images, which encode the hemodynamics extracted from GCL. Perfusion images display the phase information hidden in the eigenvectors of GCL, revealing the global phase deviation caused by the blood flow in the complicated vascular network of the imaged area.

Physiologically, the supraorbital and supratrochlear arteries dominate the blood supply to the forehead, while the facial artery predominantly supplies blood to the cheeks. The supraorbital and supratrochlear arteries are supplied by the internal carotid while the facial artery originates from the external carotid artery. Due to the difference in the travel distance and anatomical structure, it is reasonable to hypothesize that the PPG phases are different in these two regions. Our findings support this hypothesis.

We shall elaborate some technical details. In general, PPG signal is not sinusoidal and can be represented by multiple harmonics [42]; that is, we can model a clean PPG signal as 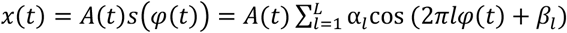, where *A* is a positive smooth function describing the amplitude modulation, *s* is a 1-periodic function describing the morphology of the nonsinusoidal oscillation, *φ*(*t*) is a strictly monotonically increasing function describing the phase of the PPG so that its derivative describes the instantaneous heart rate, *L >* 1 is the number of harmonics, and *α*_*I*_ *>* 0 and *β*_*I*_ ∈ [0,2π) come from the Fourier series of *s*. In this case, recovering the phase *φ*(*t*) from the Hilbert transform of *x*(*t*) is inaccurate due to the existence of harmonics. This situation is worsened by the existence of noise. Instead, we could better recover the phase from the fundamental component (the first harmonic) *h*_1_(*t*) ≔ *A*(*t*)*α*_1_*cos* (2π*φ*(*t*) + *β*_1_), thanks to the Nuttall theorem [43] and the assumption that *A*(*t*) in general oscillates slowly compared with the *cos* (2π*φ*(*t*) + *β*_1_). This is the main reason we design the algorithm with the first harmonic in (1). Note that the image stream is sampled at 30 Hz and we upsample the PPGI signal by a factor of 30 to reach 900 Hz. The upsampling is used to enhance the temporal resolution during the alignment. Specifically, when the sampling rate is 30Hz, the temporal resolution of alignment is 1/30 s, but after upsampling, the temporal resolution of alignment is enhanced to 1/900 s. While it is possible to use a higher upsampling rate, we found 900 Hz sufficient to achieve a reasonable result and kept to it.

Second, note that since the PPGI signals *x*_*i*_ and *x*_*j*_ are usually noisy, *c*_*i,j*_ is a noisy estimate of the global phase shift of *x*_*i*_ and *x*_*j*_. The capability of recovering phase from the usually noisy PPGI signal with the GCL-based synchronization algorithm is supported by the robustness of GCL to noise [37], [44]. In addition to the robustness of GCL, we shall also discuss the geometric meaning of GCL. Consider a simplest and ideal situation that PPGI is clean and faithfully representative of the blood flow changes, and there is no geometric constraint among pixels. In this case, *C* can be constructed without setting 0 (note that setting 0 using the geometric relationship is equivalent to applying a kernel, which helps capturing the geometry. See [36] for the analysis of GCL under the manifold setup and how it helps reconstructing the manifold.); that is, the (*i, j*)-th entry of *C* is simply *c*_*i,j*_ for any *i, j*. In this case, *C* is a rank one matrix as *C* = θθ^*^, where θ ∈ ℂ^L^ so that θ(*i*) encodes the phase of the blood flow underlying the *i*^th^ channel. In this simplest case, the top eigenvector of *C* gives the full information of phase in each channel. However, in practice the PPGI is noisy, and there are geometric constraints imposed by the vascular network. We thus only trust the phase information when two channels are close by, which leads to the construction of *C* in our algorithm. Geometrically, the top eigenvector of *C* gives us an estimate of the most synchronized phase information over the vascular network on the face. In this paper, we focus only on the top eigenvector. The higher eigenvectors of *C* encode more geometric information associated with the underlying geometric structure associated with the blood vessel distribution. We will explore this topic in our future work.

Third, the phase deviation of PPGI signals among channels could be understood as the consequence of pulse transit time caused by the complicated anatomical structure. Finding this phase relationship is itself the focus of GCL-based synchronization, where the synchronized phase could be viewed as a surrogate of pulse transit time estimate up to a global constant. Once we get the phase deviation, we could synchronize the PPGIs recorded from different channels to obtain a higher quality final non-contact PPG signal. The main reason of obtaining this higher quality non-contact PPG signal can be understood using the following example. Take *f*_1_(*t*) = *cos*(2π*t*) + *ξ*_1_(*t*) and *f*_2_(*t*) = *cos*(2π*t* + δ) + *ξ*_2_(*t*), where δ *>* 0 is a phase shift and *ξ*_1_(*t*) and *ξ*_2_(*t*) are random noises that are independent. To simplify the discussion, assume the standard deviations of *ξ*_1_(*t*) and *ξ*_2_(*t*) are both 1. By taking a direct mean without synchronization, while the noise is attenuated by the independence and taking the average, the magnitude of the signal part is also attenuated since 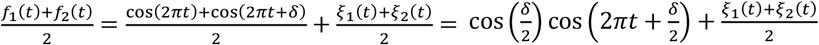 by the trigonometric identity. Clearly, the standard deviation of 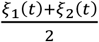 is 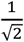, while 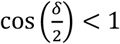 when δ *>* 0. Depending on δ, we may not obtain an enhanced signal to noise ratio. On the other hand, by eliminating the phase deviation of *f*_2_(*t*), denoted as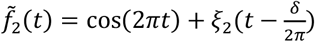, since 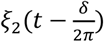is still independent of *ξ*_1_(*t*), taking a mean will attenuate the noise strength but keep the signal strength; that is, 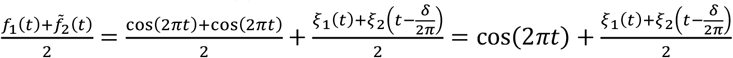, where the standard deviation of 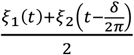 is still 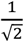, while the signal is fully recovered without attenuating the signal strength. See Fig. 1 for an illustration of this fact.

It is noteworthy to mention that the phase of PPG signals (in our case captured by the first eigenvector of GCL) has been studied in different contexts. In [12], the authors applied Discrete Fourier Transform to attempt to reveal the carotid artery or to differentiate normal skin with comparison with port wine stains. Another approach can be found in [45], where the lock-in amplification is used for phase estimation. It is worth noting that the phase shift can be calculated across different frequency bands across PPGI signals (in our case, we focus on the frequency band associated with the first harmonic), e.g. low-frequency oscillation phase distribution which shows as promising tool to study autonomic nerve system responses to external stimuli [10]. Studying and mapping the phase shift across imaged area may reveal the locations that the propagating pulse wave reached at the same time, or the positions of the subcutaneous tissue perforators. It also encodes the locations where motion or ballistocardiographic (BCG) artifacts may be present, or we may have encountered a counter-phase of the original PPGI signal [46], [47], [48]. To the best of our knowledge, perfusion maps generated using the phases of the first eigenvector of GCL have not yet been published and may provide important robust phase estimation tool which is immune to inevitable noise with theoretical support and can be used in different PPGI scenarios.

Examples of applications of our proposed signal processing technique could be used in different fields. For example measuring the body’s response to external stimuli, such as temperature (cold [10] or heat [49]) or mental stress [50]. While contact PPG is widely applied, there are situations that PPGI would be helpful. For example, it is not possible to place a contact sensor on the patient’s skin when the patient’s skin is damaged (injury, burns), or when monitoring the vital signs of premature newborns. Like the contact PPG there is the potential for the extraction of physiological parameters such as HR or blood pressure [47], [48]. Given that the dicrotic notch and its determined position may play a role in studying the vascular tone of the arterial tree our proposed algorithm shows promise in effectively capturing these features. In addition, as some studies have already pointed, the PPGI has untapped potential. Namely the challenge is to focus on truly spatial mapping of physiological parameters, especially in the context of ANS regulation or allergic reactions [10], [11], [51], [52]. There are also approaches in which the respiratory waveform is extracted from the PPGI recording. The respiratory activity can be detected using a method such as optical flow [11], [53] where our proposed algorithm could help too. Another option is to deploy this method in extracting information about SpO2, and thus in evaluation of the oxygenation level of arterial or venous or mixed blood [8], [9].

Finally, we want to summarize what our paper offers and how to use the tools. To obtain a higher quality global PPGI signal by integrating all PPGI signals from the sensed area, we use GCL for their phase alignment. This results in higher amplitude and better defined features such as dicrotic notch. If we want to work with spatial information and analyze the distribution of perfusion parameters across the imaged area, a good step here is to analyze higher eigenvectors of *C*. If we are interested in the phase relations between the PPGI waveforms, we can again use the GCL, which conveys information about the global phase of a given PPGI waveform at a given location in the image.

Here we would like to point out the limitations of this work. Although this is a small sample size, we would like to stress that this is mainly a methodological work using examples of subjects with a wide variety of characteristics. Even if the GCL is immune to noise, the quality of the PPGI signals may not always be sufficient. Instead of simple spatial averaging, other raw signal processing methods presented in the Materials and Methods should also be considered, which may offer higher SNR or robustness to motion artifacts. Moreover, capturing the subtle differences between the phase shifts given by the pulse wave propagation velocity and the geometrical dimensions of the analyzed region is challenging, especially in terms of the demands on the sampling rate (interpolation may no longer reveal and reconstruct all details) and the size of the processed data. We may need a collaborating measurement of the perfusion dynamics on the face in some form to further validate the proposed algorithm. In conclusion, despite the shortcomings, we believe that this novel approach may open the door for new applications of the PPGI.

## Data Availability

All data produced in the present study are available upon reasonable request to the authors

## Acknowledgments

The work of Stefan Borik was supported by the National Scholarship Programme of the Slovak Republic (Application No. 47518). Heartfelt thanks also go to Prof. Vladimir Blazek from RWTH Aachen, a native of the former Czechoslovakia, who is considered as the pioneer of photoplethysmography imaging and who contributed greatly to the field and the scientific and personal growth of Stefan Borik.

## Conflicts of Interests/Financial Disclosures

None

## Author contributions

SB: idea, literature review, data analysis and write-up. HTW: idea, literature review, data analysis and write-up. KHS: idea, literature review, data collection, and write-up. AAA: idea, literature review, data collection, and write-up.

## Notes

### Competing Interest Statement

The authors have declared no competing interest.

### Clinical Trial

NCT03592290

### Author Declarations

The study protocol was approved by the Institutional Review Board of Yale University (No. 2000031899). Informed consent was obtained from all subjects prior to the study. All methods were performed in accordance with the study protocol and with the Declaration of Helsinki.

### Summary of Updates

Revised version based on reviewers' critiques/comments.

